# Wastewater monitoring of SARS-CoV-2 gene for COVID-19 epidemiological surveillance in Tucumán Argentina

**DOI:** 10.1101/2023.11.22.23298851

**Authors:** María Cecilia D’Arpino, Pedro Eugenio Sineli, Gustavo Goroso, William Watanabe, María Lucila Saavedra, Elvira María Hebert, María Alejandra Martínez, Julieta Migliavaca, Silvina Gerstenfeld, Rossana Elena Chahla, Augusto Bellomio, Virginia Helena Albarracín

**Affiliations:** Laboratory of Molecular and Ultraestructural Microbiology, Centro Integral de Microscopía Electrónica,(CIME-CONICET), Universidad Nacional de Tucumán, Facultad de Agronomía, Zootecnia y Veterinaria, Tucumán, Argentina; Planta Piloto de Procesos Industriales Microbiológicos (PROIMI-CONICET), Tucumán, Argentina; Laboratorio de Processamento de Sinais e Modelagem de Sistemas Biológicos. Núcleo de Pesquisas Tecnológicas. Universidade Mogi das Cruzes, Sao Paulo, Brasil; Instituto Superior de Investigaciones Biológicas (INSIBIO, CONICET-Universidad Nacional de Tucumán), Tucumán, Argentina; Centro de Referencia para Lactobacilos (CERELA-CONICET), Tucumán, Argentina; Ministerio de Salud, Gobierno de Tucumán (SIPROSA). Tucumán, Argentina; Facultad de Ciencias Naturales e Instituto Miguel Lillo, Universidad Nacional Tucumán. Tucumán, Argentina

**Keywords:** SARS-CoV-2, Surveillance, Monitoring, Wastewater, Tucumán

## Abstract

Epidemiology based on the detection of pathogens in wastewater is extremely useful in providing information about a population’s health status. This study aimed to analyze and report the epidemiological dynamics of SARS-CoV-2 in the province of Tucumán, Argentina during the second and third surges of COVID-19 between April 2021 and March 2022. The study aimed to quantify SARS-CoV-2 RNA in wastewater, correlating it with clinically reported COVID-19 cases. Wastewater samples (n=72) were collected from 16 sampling points located in 3 cities of Tucumán (San Miguel de Tucumán, Yerba Buena y Banda del Río Salí). Detection of viral nucleocapsid markers (N1 gene) was carried out using one-step RT-qPCR. Viral loads were determined for each positive sample using a standard curve. A positive correlation (p<0.05) was observed between viral load (copies/mL) and the clinically confirmed COVID-19 cases reported during the sampling period in San Miguel de Tucumán. Our research findings provided a crucial insight into the dynamics of SARS-CoV-2 infection during epidemic outbreaks. The implementation of wastewater monitoring has proven to be an invaluable epidemiological tool, facilitating early detection of potential surges in COVID-19 cases, and enabling a comprehensive tracking of the pandemic. Our study underscores the significance of incorporating SARS-CoV-2 genome-based surveillance as a standard practice which will contribute to anticipating any future spikes in infections.

## INTRODUCTION

The Coronavirus disease 2019 (COVID-19) pandemic has caused significant public health concerns, declared a global health emergency by the World Health Organization (WHO). As of October 25, 2023, the COVID-19 pandemic has caused 771,549,718 reported cases and 6,974,473 deaths worldwide (WHO, 2023).

COVID-19 is the result of an infection with SARS-CoV-2 (SARS Coronavirus 2), an enveloped, positive-sense single-stranded RNA virus belonging to the Order Nidovirales, Family Coronaviridae, genus Betacoronavirus (Alexandersen et al., 2020; Kadam et al., 2021; Shereen et al., 2020; Zhou et al., 2020; Zhu et al., 2020).

Human-to-human transmission occurs through droplets, aerosols, and contaminated surfaces originating from an infected person’s breath secretions (Jones et al., 2020; Sharma et al., 2021). Patients infected with SARS-CoV-2 exhibit diverse symptoms, including fever, cough, shortness of breath, sore throat, and headaches. Clinical manifestations vary from asymptomatic infection or mild respiratory disease to severe pneumonia, respiratory failure, and even multi-organ failure (Guan et al., 2020; Jiang et al., 2020). Gastrointestinal symptoms, such as nausea, vomiting, loss of appetite, diarrhoea, and abdominal pain, have also been reported (Adhikari et al., 2022; Kariyawasam et al., 2021; Megyeri et al., 2021). Notably, the gastrointestinal tract contains a substantial amount of angiotensin-converting enzyme 2 (ACE-2), a receptor used by SARS-CoV-2 for cell entry (Hamming et al., 2004; Lee et al., 2020; Lewandowski et al., 2023). This allows the virus to infect human enterocytes in the gastrointestinal tract, leading to viral replication and excretion in faeces (Jones et al., 2020). Therefore, SARS-CoV-2 RNA is commonly detected in wastewater, with concentrations of up to 107 copies per g (Ahmed et al., 2020a; Thompson et al., 2020).

Wastewater-based epidemiology (WBE) is a methodology that allows an assessment of the population’s exposure to infectious agents such as SARS-CoV-2. WBE offers valuable information on the temporal and spatial dynamics of population exposure. It provides information on a community’s health status concerning etiological agents found mainly in human faeces or urine. (Ahmed et al., 2021; Giraud-Billoud, 2021; Prado et al., 2021; Randazzo et al., 2020).

Given the magnitude of the COVID-19 crisis, there is a pressing need for integrated alert and surveillance systems to detect and model SARS-CoV-2 infections. Accurate assessment of the number of infected individuals is challenging, especially due to a significant proportion of carriers displaying mild or no symptoms but capable of excreting and transmitting the virus (Maréchal et al., 2023; Rothe et al., 2020). Sewage surveillance can monitor virus circulation in the community, covering both symptomatic and asymptomatic carriers. Moreover, WBE can serve as an early warning system for SARS-CoV-2, outbreak detection, and identification of circulating strains (Li et al., 2023). Furthermore, the use of WBE for the detection of SARS-CoV-2 RNA in wastewater may provide early indicators of an increase in clinical cases at the population level (Ahmed et al., 2020b; Ahmed et al., 2021; Larsen and Wigginton, 2020; Medema et al., 2020).

This study represents the first attempt to monitor SARS-CoV-2 epidemiological evolution using WBE analysis in Tucumán, Argentina. The study involved the analysis of wastewater samples collected during the COVID-19 pandemic between April 2021 and March 2022 from various sources, including manholes, wastewater treatment plants, and hospital effluent exits. These samples were subjected to viral concentration and RNA isolation methods, followed by quantification of SARS-CoV-2 nucleocapsid markers by RT-qPCR. Finally, this study correlated the number of genetic copies of the SARS-CoV-2 N1 gene in wastewater with reported COVID-19 cases.

## MATERIALS AND METHODS

### 2.1 Wastewater sampling

Tucumán province is in the Northwest of Argentina with a population of 1.703.186 inhabitants (http://estadistica.tucuman.gov.ar/index.php/censo-2022). Seventy-two wastewater samples were collected between April 2021 and March 2022. The sampling period covered the 2^nd^ and 3^rd^ wave of COVID-19 pandemic in Tucumán. The samples were collected from various sources: manholes located in San Miguel de Tucumán (n9) and Yerba Buena (n4), a wastewater treatment plant (WWTP), (n1) located in San Miguel of Tucumán and the wastewater inspection chambers of two hospitals, one located in San Miguel de Tucumán (n1) and the other in Banda del Río Salí (n1). All samples were taken early in the morning by collecting 500 mL of water in sterilized plastic containers. The collected samples were brought on ice to the laboratory for processing within 24 hours. No precipitation or storms occurred on or before the sampling days.

### 2.2. PEG concentration method

The samples were thermally inactivated to increase the safety of their handling (Hasan et al., 2021; Pastorino et al., 2020). PEG-based viral concentration method was used, which according to the literature is one of the most efficient methods in terms of viral recovery (Barril et al., 2021; Iwai et al., 2009; Thongprachum et al., 2018). 200 mL was transferred to a sterile glass bottle with a screw cap containing 20 g of 8% (w/v) polyethylene glycol (PEG) 6000 (Sigma Aldrich SRL, Argentina) and 4.5 g of 0.2 M (p/v) (Cicarelli). It was shaken to homogenize, and the mixture was allowed to stand overnight at 4°C for viral precipitation. Viruses were concentrated by two-step centrifugation, 7670 g for 10 min and then 7990 g for 1 h at 4°C. The pellet was suspended in 1 mL of TRI Reagent® (Molecular Research Center. Inc) to obtain the homogenate) and stored at −70°C.

### 2.3. SARS-CoV-2 RNA purification

Viral RNA was extracted from TRI Reagent® (Molecular Research Center) homogenate and purified by using a Puro Virus RNA (Productos Bio-Lógicos, Argentina), according to the manufacturer’s instructions. Briefly, 140 µl of the TRI Reagent homogenate was mixed with 560 µl of lysis buffer A plus ARN Carrier (10,7 ug/mL) and subjected to pulse-vortexing for 15 seg and incubated for 15 min at room temperature. Then, 560 µl of cold 100% ethanol was added and incubated for 5 min at room temperature. The lysate was transferred to the column and centrifuged at 6000 g for 1 min. Then was washed with buffer w1, centrifuged at 6000 g for 1 min, and continued with two washes with buffer w2. The column was then dried by centrifuging at 12,000 g for 3 min and then RNA elution was performed in 50 µl of RNase-free water by centrifugation at 12,000 g for 1 min. Purified RNA was stored at −80 °C. The concentration and quality of RNA were measured using a µDrop Plate (Thermo Scientific).

### 2.4. Detection of SARS-CoV-2 genetic marker in wastewater samples

Detection of SARS-CoV-2 genetic marker from wastewater was carried out by one-step RT-qPCR. The N-gene encoding the viral nucleocapsid (N) was amplified as target using the primers 2019-nCoV_N1F: GACCCCAAAATCAGCGAAAT, 2019-nCoV_N1R: TCTGGTTACTGCCAGTTGAATCTG and the probe was, 2019-nCoV_N1P: FAM-ACCCCGCATTACGTTTGGTGGACC-BHQ1, validated by the US Centers for Disease Control and Prevention (CDC, 2020) (Lednicky et al., 2020). RT-qPCR mix was prepared using qScript XLT 1-Step RT-PCR (Quantabio) in a one-step system. Reaction mix (10 µl) consisted of One Step RT master Mix, Qscript ROX Reference Dye, 0.50 pmol/mL REV primer, 0.5 pmol/mL FW primer and 0.25 pmol/mL FAM labeled TaqMan probe, nuclease-free water. A real-time (RT-qPCR) assay was performed in a StepOnePlus™ Real-Time PCR System (Applied Biosystems). The RT qPCR program set up was, one cycle for retrotranscription at 50°C for 10 min, then a preheating at 95 °C for 1min and 45 cycles of amplification at 95 °C for 10 s, and 60 °C for 1 min. Each RNA was analyzed in duplicate, and every RT-qPCR assay included negative (nuclease-free water) and positive controls.

### 2.5. Quantification of SARS-CoV-2 in wastewater samples

The viral load in those wastewater samples that tested positive for SARS-CoV-2 RNA was quantified using a standard curve of a plasmid containing the SARS-CoV-2 N gene (2019-nCoV_N_Positive Control, IDT).Linear dynamic range was evaluated among 50 and 200000 genetic copies. The standard curve used had an R^2^ value of 0.995 (slope = −3.5167; y-intercept = 32.86). The results were expressed as the number of genetic copies in 1 mL of wastewater (copies/mL).

### 2.6. Statistical analysis

Pearson’s correlations analyses were performed to test the correlation between COVID-19 cases and the number of genetic copies by 1 mL of wastewater samples. The significance level was fixed at *p*< 0.05.

### 2.7. Variables related to epidemiological surveillance

All epidemiological data covering the period from April 2021 to March 2022, considered in this investigation, were provided by the environmental department of the Ministry of Health of Tucumán.

## RESULTS

### 3.1. Epidemiological data in the Tucumán province, Argentina

The evolution of daily COVID-19 cases in Tucumán during the first, second, and third waves is shown (Figure 1). There were over 90% of cases reported in the metropolitan area of Tucumán, which covers four different districts of the province: Capital San Miguel de Tucumán, Cruz Alta, Yerba Buena, Tafí Viejo and Lules. The first wave occurred during the second semester of 2020: the second week of August to the end of November with a peak on October 15, where approx. 1300 daily cases were recorded. In 2021, a second wave lasted 24 weeks, from March to August. Daily cases increased from 166 in early March to 755 in mid-April. Movement restrictions were imposed on April 8, followed by a confinement declaration on May 22. June showed a peak with up to 1400 cases/day, with 155345 confirmed cases in the province as of June 28, 2021. 65% of these cases were concentrated in San Miguel de Tucumán. A significant decrease in cases occurred from August, with fewer than 50 confirmed daily cases until December. A subsequent increase in cases was observed during the third wave, which extended from mid-December to mid-February. On January 10, 6500 positive cases were recorded. By March 2022, cases had dropped to five per day.

**Figure 1:**
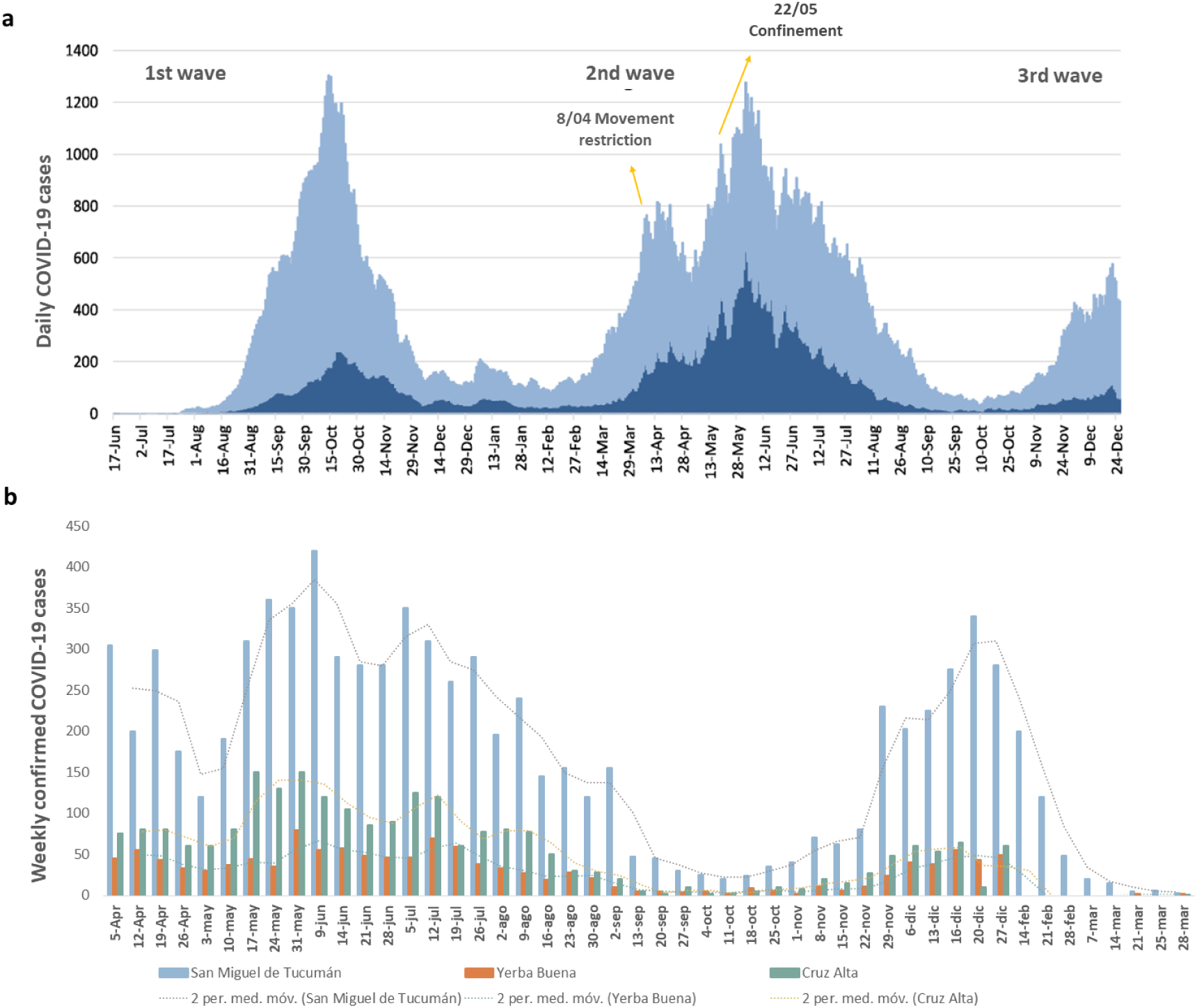
Epidemiological data of the SARS-CoV-2 pandemic in Tucumán. a, The curve of confirmed cases of Covid-19 in Greater San Miguel de Tucumán (light blue) and the interior of the province (dark blue) from June 2020 to March 2022. b, Temporal evolution of the weekly number of COVID-19 cases during the second and third waves in San Miguel de Tucumán, Yerba Buena, and Cruz Alta.

### 3.2 SARS-CoV-2 genetic marker detection in wastewater

Taking into account the above-referred epidemiological context, wastewater sampling was conducted during the second and third waves; more specifically in April, June, September, December (2021) and March 2022. In total, 13 manholes from the cities with the most reported cases were sampled: San Miguel de Tucumán (n9) and Yerba Buena (n4). In addition, the main wastewater treatment plant of the metropolitan area, San Felipe, was sampled for a general reference together with two COVID19-reference hospitals’ sewage systems (positive controls): i) Zenón J. Santillán Health Center (San Miguel de Tucumán); ii) Eva Perón Hospital (Banda del Rio Salí, Cruz Alta). Figure 2.a shows the geolocation of each sampling point (Table 1).

**Figure 2:**
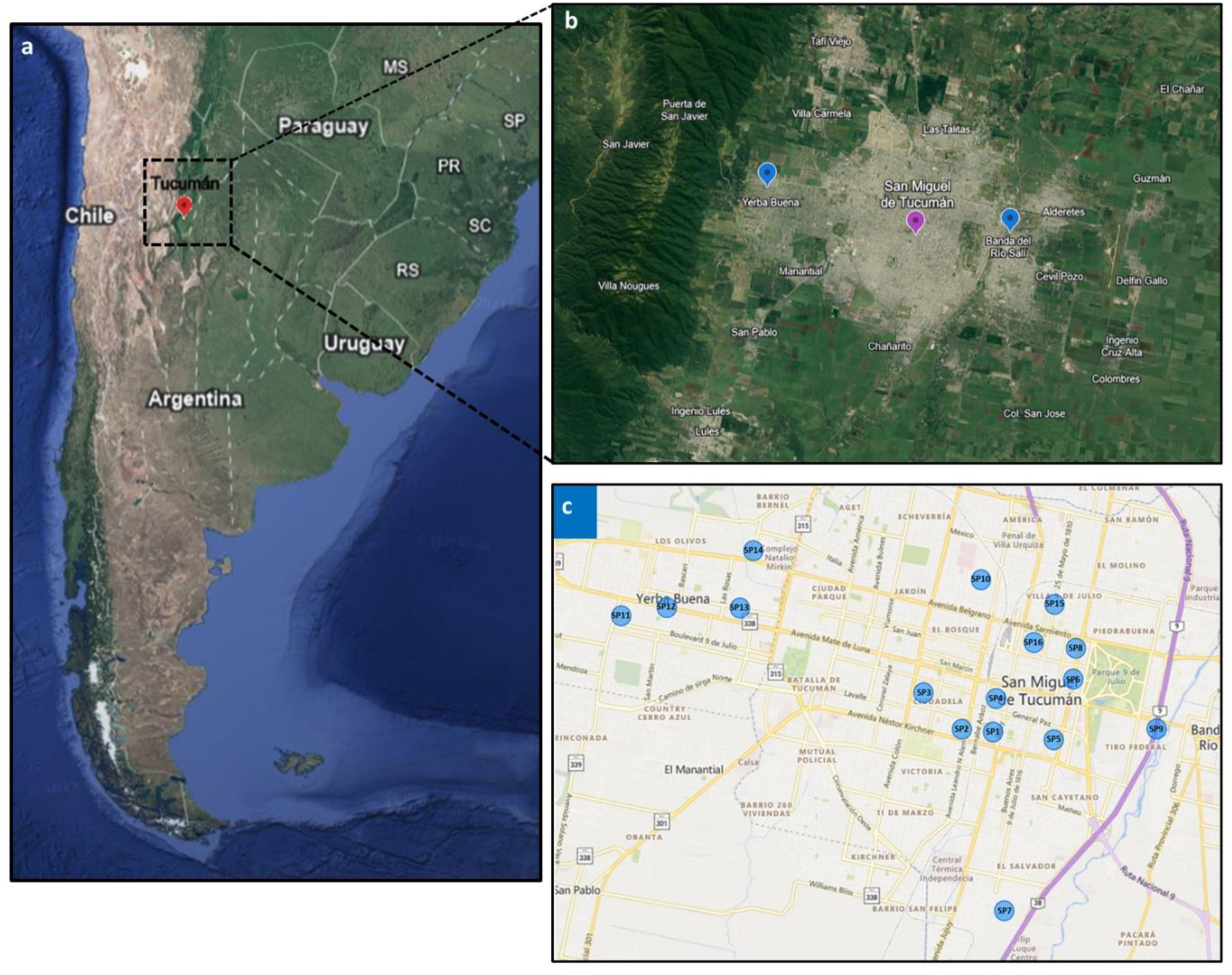
Sampling areas of the province of Tucumán, Argentina. The dotted line (a) shows the location of Tucumán. In b the markers indicate the cities in which sampling was carried out: San Miguel de Tucumán, Yerba Buena, and Banda del Río Salí. c the visualization of the different sampling points. The light blue circles show the location of each of the sites from which the samples were obtained.

**Table 1:** Location and origin of the sampling points. Coordinates of each of the points, and the origin of the samples is also indicated. SP (Sample Point). SP 1, SP2, SP3, SP4, SP5, SP6, SP10, SP15, SP16 correspond to manholes in San Miguel de Tucumán. SP11, SP12, SP13, and SP14 correspond to Yerba Buena manholes. SP7 corresponds to the San Felipe sewage treatment plant entrance. SP 8 corresponds to the Health Center Zenon J. Santillan wastewater inspection chamber in San Miguel de Tucumán, and SP9 Eva Peron Hospital wastewater inspection chamber in the city of Banda del Rio Salí (Cruz Alta).

| ID | City | Coordinates | Source |
| --- | --- | --- | --- |
| SP01 | SM. Tucumán | -26.8388842-65.2148118 | Manhole |
| SP02 | SM. Tucumán | -26.8383123-65.2220305 | Manhole |
| SP03 | SM. Tucumán | -26.8309718-65.2309513 | Manhole |
| SP04 | SM. Tucumán | -26.8321377-65.2140929 | Manhole |
| SP05 | SM. Tucumán | -26.8404344-65.2007152 | Manhole |
| SP06 | SM. Tucumán | -26.828233-65.1961796 | Manhole |
| SP10 | SM. Tucumán | -26.8082848-65.2175903 | Manhole |
| SP11 | Yerba Buena | -26.8155347-65.3011424 | Manhole |
| SP12 | Yerba Buena | -26.8138829-65.2905533 | Manhole |
| SP13 | Yerba Buena | -26.8139626-65.2735795 | Manhole |
| SP14 | Yerba Buena | -26.8024385-65.2704578 | Manhole |
| SP15 | SM. Tucumán | -26.8132221-65.2005151 | Manhole |
| SP16 | SM. Tucumán | -26.8208967-65.2053334 | Manhole |
| SP07 | SM. Tucumán | -26.87476172-65.2122031 | San Felipe sewage treatment plant entrance |
| SP08 | SM. Tucumán | -26.82204-65.1955586 | Health Center Zenon J. Santillan wastewater inspection chamber |
| SP09 | Banda del R. Salí | -26.8382557-65.1768163 | Eva Peron Hospital wastewater inspection chamber |

Over time, we analysed 72 samples and detected SARS-CoV-2 RNA via RT-qPCR. The average Cq values collected from manhole wastewater samples were as follows: April 2021 (29.73 ± 4.55), June 2021 (32.78 ± 1.73), September 2021 (41.24 ± 0.51), and December 2021 (39.02 ± 4.11) (mean ± SEM). The Cq values correspond to the San Felipe wastewater treatment plant (SP7), and the wastewater inspection chamber of the Centro de Salud Zenón J. Santillán (SP8) and Eva Perón (SP9) hospitals (supplementary material).

All sampling points, except SP1 and SP2, showed evidence of SARS-CoV-2 at least once during the sampling period. Specifically, SP3 exhibited a low viral load in September 2021. Additionally, sample points 12, 14, and 15 were only detected in June. Table 2 summarizes RNA viral load data for each sampling point during the second wave. In April 2021, 6 out of 16 samples tested positive. The highest viral load was observed at SP4. June 2021 sampling, coinciding with the second wave, yielded 9 positive samples out of 16. In September 2021, 5 of 8 points analysed in San Miguel de Tucumán turned positive. In December 2021, 10 out of 16 samples tested positive. On the other hand, sites SP5, SP11, and SP16 only showed virus presence in December. No viral RNA was detected in the March 2022 samples.

**Table 2:** Sars-CoV-2 viral load. Viral load calculated for each sampling point during the months of April, June, September, and December 2021 and March 2022.

| ID | Load viral (copies/mL) |  |  |  |  |
| --- | --- | --- | --- | --- | --- |
|  | Apr-21 | Jun-21 | Sep-21 | Dic-21 | Mar-22 |
| SP1 | ND | ND | ND | ND | ND |
| SP2 | ND | ND | ND | ND | ND |
| SP3 | ND | ND | 0,64 ± 0,42 | ND | ND |
| SP4 | 250142.49 ± 66554.29 | 3229.37 ± 974.32 | 5.60 ± 1.34 | 10.48 ± 2.79 | ND |
| SP5 | ND | ND | ND | 3096.75 ± 201.14 | ND |
| SP6 | ND | ND | 5.12 ± 4.11 | 64.26 ± 17.67 | ND |
| SP7 | 1451.06 ± 486.40 | 5334.32 ± 1703.69 | 12.54 ± 5.46 | 1.13 ± 0.51 | ND |
| SP8 | 5222.14 ± 777.78 | 32725.17 ± 13010.01 | 28.86 ± 3.34 | 12260.28 ± 966.64 | ND |
| SP9 | 333.06 ± 0.77 | 6041.19 ± 2141.08 | ... | 1.78 ± 0.57 | ND |
| SP10 | 1066.21 ± 375.09 | 2708.26 ± 720.57 | ... | 28.79 ± 3.73 | ND |
| SP11 | ND | ND | ... | 2.50 ± 1.14 | ND |
| SP12 | ND | 441.00 ± 95.47 | ... | ND | ND |
| SP13 | 1966.81 ± 722.67 | 2228.73 ± 999.08 | ... | 4.99 ± 5.14 | ND |
| SP14 | ND | 196.11 ± 43.35 | ... | ND | ND |
| SP15 | ND | 1178.28 ± 376.32 | ... | ND | ND |
| SP16 | ND | ND | ... | 1.17 ± 0.85 | ND |

### 3.3 Relationship between SARS-CoV-2 viral load and cases of Covid 19 in Tucumán

We analysed the relationship between the SARS-CoV-2 viral loads and the number of cases based on the detection of the N1 marker in the COVID-19 cases registered in each city during the second and third emergences of COVID-19 to obtain a global perspective of the viral load and the number of cases. Among the points that have shown detectable viral load over time (Figure 3), are SP4, SP7, and SP8 from San Miguel de Tucumán, SP10 and SP13 in Yerba Buena, and SP9 in Banda del Ro Sal. The viral load obtained for sampling points SP4, SP7, and SP8 were related to the positive cases of COVID-19 registered in San Miguel de Tucumán on the sampling day during the months of analysis. Figure3.a allows us to observe a high viral load in SP4 at the beginning of sampling (250142.49 copies/mL). On the other hand, in June, when positive cases peaked, a viral load of 3229.37 copies/mL was detected. Then in September, the viral load recorded was very low, 5.6 copies/mL, and remained at low levels in December, when an increase in positive cases was recorded (10.48 copies/mL). In March, no viral load was detected. Figure 3.b shows the relationship between the SARS-CoV-2 viral load detected at the entrance to the San Felipe wastewater treatment plant (SP7) with positive COVID-19 cases. In this scenario, we observed an initial viral load of 1451.06 copies/mL in April, which subsequently increased to 5334.32 copies/mL by June, coinciding with an increase in positive cases. In September it dropped to 12.54 copies/mL and remained very low during December at 1.13 copies/mL, becoming undetectable in March 2022. Figure 3.c shows the relationship of the viral load with positive cases at The Health Center (Zenon J. Santillán). The viral load in the April sample was 5222.14 copies/mL, experiencing a substantial surge to 32725.17 copies/mL in June. Subsequently, in September, the recorded viral load markedly decreased to a mere 28.86 copies/mL, only to rise again to 12260.28 copies/mL in December. The situation took a positive turn by March 2022, as no viral RNA was detected at that point. We performed a correlation analysis between COVID-19 cases and the load of the SARS-CoV-2 N1 marker in the samples. The Spearman test showed values of rs = 0.9, p (2-tailed) = 0.03739, evidencing a statistically significant association for both variables. Figure 3.d shows the relationship between the SARS-CoV-2 viral load in SP9 wastewater, and the COVID-19 cases recorded in Cruz Alta during the test months. In April, the viral load was 333.06 copies/mL, in June a notable increase was observed to 6041.19 copies/mL, while in December a sharp decrease was recorded at 1.78 copies/mL, becoming undetectable in March 2022. Figure 3.e enables us to examine the correlation between viral load and positive cases at SP10 and SP13 points, which are associated with manholes situated in the city of Yerba Buena. The patterns observed for both were quite alike; consequently, the viral loads documented in April were 1066.21 copies/mL for SP10 and 1966.81 copies/mL for SP13, respectively. In June, at both points an increase was detected to 2708.26 and 2228.73 copies/mL for SP10 and SP13 respectively, coinciding with an increase in the number of confirmed positive cases for COVID-19. In September, a notable decrease was recorded at both points 28.79 and 4.99 copies/mL for SP10 and SP13 respectively. Finally, in March 2022, no N1 marker of SARS-CoV-2 was detected.

**Figure 3:**
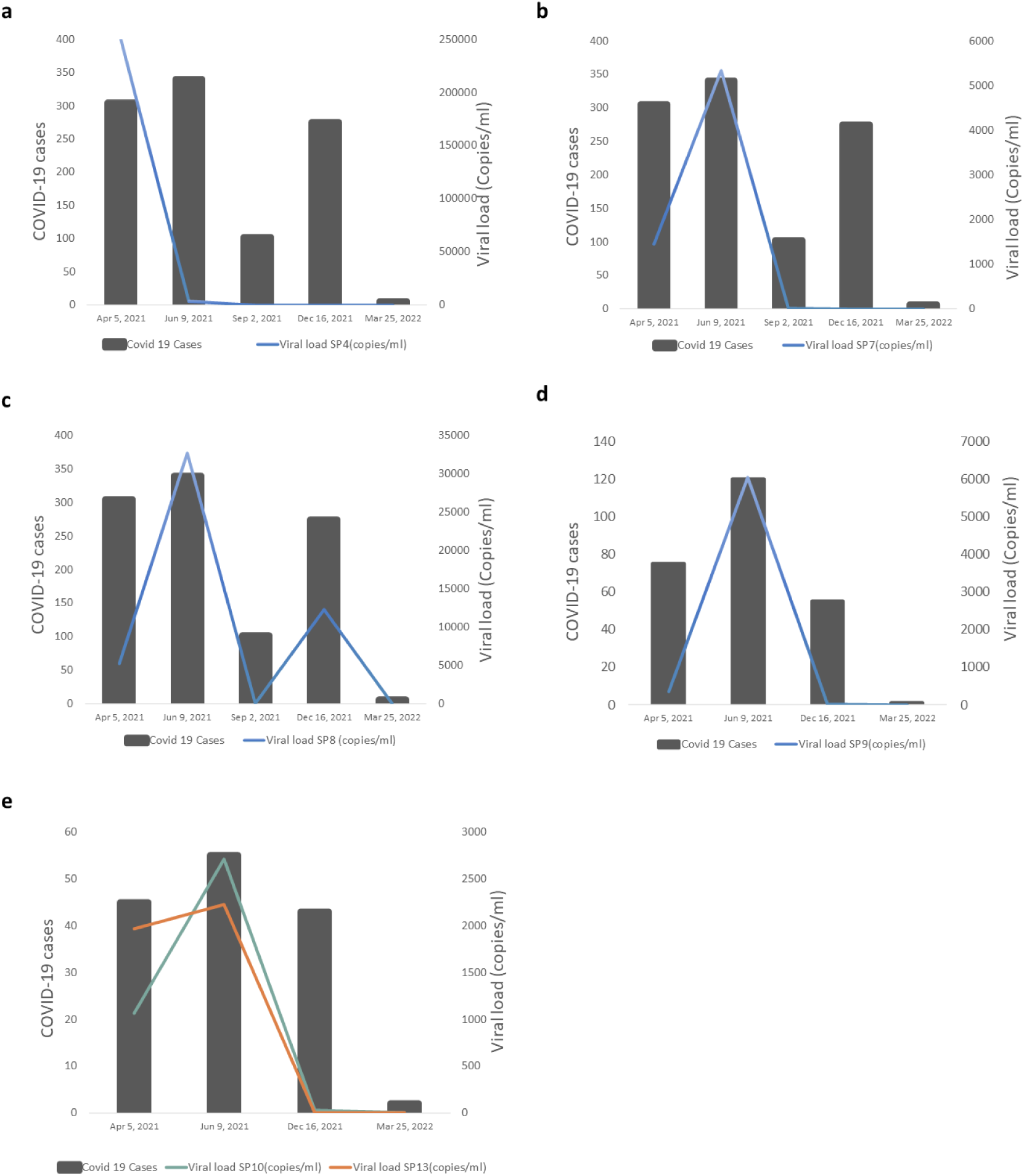
Relationship between Sars-CoV-2 viral load and positive COVID-19 cases. a, in samples obtained from sewer SP4 in San Miguel de Tucumán; b, in samples taken from the San Felipe sewage treatment plant entrance (SP7); c, samples collected from inspection chamber of the Hospital Centro de Salud (Zenón J. Santillán) (SP8). a, b, and c correspond to points located in San Miguel de Tucumán sampled from April 5, 2021, to March 25, 2022. d, Viral load of Sars-CoV-2 in wastewater obtained from Eva Peron Hospital wastewater inspection chamber (SP9), this point is in Banda del Río Salí, Cruz Alta and was sampled from April 5, 2021, to March 25, 2022. e, Viral load of Sars-CoV-2 in wastewater obtained from sewers SP10 and SP13, both located in Yerba Buena. Samples were collected from April 5, 2021, to March 25, 2022.

## DISCUSSION

In Argentina, from 3 January 2020 to 8:20 am CEST, 19 July 2023, there have been 10,044,957 confirmed cases of COVID-19 with 130,472 deaths, reported to WHO. As of 7 October 2022, a total of 109,652,736 vaccine doses have been administered. The first wave lasted 12 weeks with a peak in October 2020, while the second wave lasted about 24 weeks with an epidemic peak in June 2021. In this work, sampling was carried out in April, June, September, December 2021, and March 2022. It is important to note that in 2021, on April 8, a movement restriction was established since the second surge of COVID-19. As of June 28, 2021, confirmed cases in the province of Tucumán amounted to 155,345 from 591,339 studied samples. The cumulative index per 10,000 inhabitants was 916.7. During the second wave, 65% of the cases in the entire province correspond to San Miguel de Tucumán.

Detection of SARS-CoV-2 RNA in wastewater has been reported worldwide (Ahmed et al., 2020; La Rosa et al., 2020; Wani et al., 2023;Wu et al., 2020). These studies show the usefulness of monitoring viral RNA in wastewater for surveillance of SARS-CoV-2 infection at the level of the entire population. SARS-CoV-2 monitoring in the WBE is based on the excretion of the virus through the gastrointestinal system. In this way, viral genetic material can be detected in drainage as well as in wastewater treatment plants (Ahmed et al., 2021; Atoui et al., 2023), even before COVID-19 patients are detected in the population and up to 21 days after the number of diagnosed cases decreases to baseline (Amirian, 2020; Yeo et al., 2020). The present article is part of a federal research program aimed at examining wastewater to evaluate the circulation of the SARS-CoV-2 virus in Argentina during the pandemic. Theinvestigationsprovidedepidemiological data concerningdifferentprovinces; Buenos Aires (Barrios et al., 2021; Iglesias et al., 2021), Santa Fé (Reno et al., 2022), Mendoza (Giraud-Billoud, et al., 2021), Córdoba (Masachessi et al., 2022), Salta (Cruz et al., 2023; Maidana-Kulesza, et al., 2022). It is the first surveillance report carried out in the Province of Tucumán to develop a strategy for detecting SARS-CoV-2 at the city level from wastewater. Viruses must be recovered from these samples. Different strategies are used, including precipitation with (PEG) for viral concentration (Ahmed et al., 2020a; Wu et al., 2020), of which there are variants. In this work, SARS-CoV-2 was concentrated from wastewater by PEG precipitation. These approaches have already been used to recover SARS-CoV-2 from environmental water samples (Barril et al., 2021; La Rosa et al., 2020; Pérez-Cataluña et al., 2021; Randazzo et al., 2020) because they demonstrated adequate viral recoveries when wastewater matrices were artificially seeded with SARS-CoV-2 (Ahmed et al., 2020a; Barril et al., 2021; Lewis and Metcalf, 1988). In addition, it has been reported as a method that allows satisfactory recovery at a low cost (Wani et al., 2023). We detected SARS-CoV-2 RNA by RT-qPCR using N1 primer sets, already used and validated for clinical laboratory testing. In this work, the first sample was taken on April 5, during the beginning of the second wave of COVID-19. Although 6 of the 16 spots were positive, in SP4 we detected a large increase in the N1 marker for SARS-CoV-2 (copies/mL), a few days before COVID-19 cases increased (Table 2). At this time, in Argentina, the movement restriction was established due to a sustained increase in the curve of daily cases during the last four weeks in the city of Buenos Aires, and in nine provinces including Tucumán (Directorate of Epidemiology of the Ministry of Health of the Nation, 2021). In a study conducted in Santa Fé, Argentina, increases in genetic markers N1 and N2 in wastewater treatment plants occurred 3 to 6 days before the weekly increase in COVID cases (Giraud-Billoud, et al., 2021). In New Haven, Connecticut, USA, Peccia et al. (2020) demonstrated that SARS-CoV-2 RNA concentrations from primary sewage sludge were 1 to 4 days earlier than local hospital admissions and 6 to 8 days earlier than positive SARS-CoV-2 tests.Coinciding with the increase in the number of cases in June, when the peak of the second wave occurred, we recorded that a greater number of sampling points were positive compared to April (Table 2). Our study describes low Ct values and high viral load (copies/mL) in both April and June 2021. A positive correlation between SARS-CoV-2 viral load and its transmissibility has been reported (Dadras et al., 2022). In Tucumán, a peak of positive cases of Covid-19 was registered in June. Furthermore, at this time a mortality rate per million inhabitants of 10.0 was recorded. COVID-19 patients who have been treated in the intensive care unit with severe illness have been shown to have a relatively higher viral load (Abdulrahman et al., 2021). A study conducted in large hospitals suggested that a high viral load is associated with an increased risk of death (El Zein et al., 2021). Although there are no studies in our province that report the viral load of patients during the second wave, our results could be associated with the fact that many COVID-19-positive people have a high viral load. Indeed, people infected with SARS-CoV-2 have been reported to shed the virus in feces for prolonged periods and with viral loads that can reach up to 1 × 10^6^ genomic copies (GC) per gram of fecal material (Amirian, 2020; Yeo et al., 2020). In September, positive sampling points showed a low viral load, which may be related to the decrease in the number of cases, as well as the increase in vaccination coverage. In Argentina, a decrease in the number of positive cases of COVID-19 has been observed since the beginning of August (data repository of the Center for Systems Science and Engineering (CSSE) at Johns Hopkins University). A similar pattern could be observed in Tucumán. In mid-August 2021, the percentage of vaccination coverage in both Argentina and Tucumán was 30% and in mid-December, it was 70%, while as of March 27, 2022, it reached 80% (Johns University Data Repository Hopkins Center for Systems Sciences and Engineering (CSSE)).According to CSSE, data repository at Johns Hopkins University, the increase in the percentage of the population vaccinated with full doses was associated with a decrease in the number of cases as well as a lower mortality rate evidencing the end of the second wave epidemic outbreak. Importantly, the protection of various vaccines against hospitalization and death from severe COVID-19 disease slowly decreased after a two-dose schedule of a global COVID-19 vaccine (Zhou et al., 2022). In December, 10 of the points analyzed were positive but with a very low viral load except for point SP8 corresponding to the Health Center Hospital (Table 2), possibly in relation to many people admitted to the COVID Unit. A few weeks later, the number of cases increased, leading to the third wave, with a peak of positive cases in January. During this period, a mortality rate per million inhabitants of 5.0 was reported with a vaccination coverage percentage of 80% at both the national and provincial levels. This increase in both the number of cases and mortality may be related to the circulation of the new variant B.1.1.529 “Omicron” (WHO, 2021). A decrease in the number of cases was then observed nationally (Johns Hopkins University Center for Systems Science and Engineering (CSSE) data repository). In Tucumán, at the beginning of March 2022, between 30 and 45 cases per day were recorded, while towards the end of this month, they did not exceed 5 positive cases per day. Our analysis showed that all sampling points were undetectable for the SARS-CoV-2 N1 marker. The limit of detection (LOD) of 2019-nCoV plasmid (CDC, USA) by RT-qPCR for N1 and N2 is 25 copies/reaction.

When we analyze the positive sampling points in relation to the number of cases, we find a positive correlation for points SP4, SP7, and SP8 in the months of May and June. However, at other points, it was not possible. Many authors consider that the SARS-CoV-2 RNA content in wastewater is not necessarily proportional to the changes in confirmed cases (Ai et al., 2021; Faladori et al., 2020; Wu et al., 2020).Finally, the decrease in the detection of SARS-CoV-2 positive samples in wastewater coincides with the decrease in confirmed clinical cases of COVID-19 that marks the end of the third wave of the COVID-19 epidemic.

Recent research considers that wastewater surveillance is essential for tracking the spread of SARS-CoV-2 among populations(Jiang et al., 2023; Pipes et al., 2022). Considering that wastewater represents a composite biological sample of the community, continuous monitoring could be increasingly useful in areas where clinical trial rates decline or resources for clinical trials are limited (Hoar et al., 2022). Along these lines, this work could contribute to building a WBE network with the aim of monitoring SARS-CoV-2 RNA in wastewater during the post-pandemic.

## CONCLUDING REMARKS AND PROSPECTS

Our study spanned April 2021 to March 2022, covering the second and third waves of COVID-19 in Tucumán. During this time, wastewater samples were collected from various sources, including manholes, a wastewater treatment plant, and hospital effluent exits. Positive correlations were observed between SARS-CoV-2 viral load in wastewater and reported COVID-19 cases at specific sampling points (SP4, SP7, SP8, SP9, SP10, SP13). Notably, high viral loads coincided with peaks in COVID-19 cases, providing a potential indicator for epidemiological trends.

The study identified seasonal variations in SARS-CoV-2 RNA in wastewater, with higher viral loads during peak infection periods. Moreover, lower viral loads in September 2021 aligned with decreased cases and increased vaccination coverage, highlighting the potential impact of vaccination efforts. Thus, our study emphasized the utility of WBE for early detection of SARS-CoV-2 and monitoring its circulation. The undetectability of SARS-CoV-2 RNA in wastewater samples in March 2022 corresponded with the decline in clinical cases, supporting the value of wastewater surveillance in post-pandemic monitoring.

This study represents the first attempt to monitor SARS-CoV-2 epidemiological evolution in Tucumán using WBE. Positive correlations between viral load and reported cases suggest the potential use of wastewater analysis as an early warning system. The findings underscore the importance of integrated alert and surveillance systems, especially in regions with varying vaccination coverage and evolving COVID-19 waves. In addition, our research contributes to the establishment of a WBE network for post-pandemic monitoring. Further investigations are currently in progress to unveil the dynamics of viral shedding in wastewater, considering variations in clinical cases, vaccination status, and the emergence of new variants. The expected results will have implications for public health strategies and the development of targeted interventions based on wastewater surveillance data in Tucumán, Argentina.

## 1. Credit authorship contribution statement

VHA, AB, MLS, EMH, and GG had the original research idea and designed the project. MCD, PES, AB, and MLS designed and performed the experiments. JM, SG and REC designed and performed the sampling and provided the epidemiological data. MCD, PES, WW and GG analysed the experimental and epidemiological data and produced the figures and tables. VHA, AB, MLS, EMH and GG obtained and executed funding for the original project idea. VHA, AB, MAM and REC provided instrumentation, consumables, logistics and infrastructure for the execution of the experiments. MCD, PES and VHA wrote the paper. AB, MLS, EMH, GG and MAM edited and critically analysed the manuscript. All authors read and approved the final version of the manuscript.

## Data Availability

All data produced in the present work are contained in the manuscript

## 2. Acknowledgements

The authors acknowledge the generous financial support by the National Council of Research and Technology (CONICET) and Ministry of Science, Technology and Innovation of Argentina (MINCyT) through “PROGRAMA DE ARTICULACIÓN Y FORTALECIMIENTO FEDERAL DE LAS CAPACIDADES EN CIENCIA Y TECNOLOGÍA COVID-19”. This work was funding by the project TUC-03-COVID19 “*COVID-U-MAP”*, included in the Bank of Technological and Social Development Projects of CONICET (PDTS). Our group was part of the “Federal Network for COVID detection in the environment”, Unidad Coronavirus, MINCyT. The project received financial support of the Provincial Health Ministryin the frame of COVID19 Research Program, processed by a specific agreement between the Superior Government of the Province of Tucumán, the National University of Tucumán and CONICET. MCD had a postdoctoral research scholarship from CONICET. VHA, AB, MLS, EMH, PES, and MAM are CONICET researchers.

## Abbreviations

COVID-19: Coronavirus disease-19
Cq: quantification cycle
PEG: polyethyleneglycol
SARS-CoV-2: acute respiratory syndrome-coronavirus 2
WBE: wastewater-based epidemiology
WWTP: wastewater treatment plant

