## Supplementary material for "Wastewater monitoring of SARS-CoV-2 gene for COVID-19 epidemiological surveillance in Tucumán Argentina": Supplemetary 1 and 2

| **Sample** | **Cq 1** | **Cq 2** | **Average Cq** | **SD** | **VL 1** | **VL 2** | **Average VL** | **SD** |
| --- | --- | --- | --- | --- | --- | --- | --- | --- |
| 1 | ND | ND | - | - | - | - | - | - |
| 2 | ND | ND | - | - | - | - | - | - |
| 3 | ND | ND | - | - | - | - | - | - |
| 4 | 24,7 | 24,2 | 24,5 | 0,410 | 203081,51 | 297203,48 | 250142,49 | 66554,29 |
| 5 | ND | ND | - | - | - | - | - | - |
| 6 | ND | ND | - | - | - | - | - | - |
| 7 | 31,9 | 32,7 | 32,3 | 0,520 | 1795,00 | 1107,13 | 1451,06 | 486,40 |
| 8 | 30,2 | 30,5 | 30,3 | 0,228 | 5772,11 | 4672,16 | 5222,14 | 777,78 |
| 9 | 34,5 | 34,5 | 34,5 | 0,004 | 333,61 | 332,52 | 333,06 | 0,77 |
| 10 | 33,2 | 32,4 | 32,8 | 0,547 | 800,98 | 1331,44 | 1066,21 | 375,09 |
| 11 | ND | ND | - | - | - | - | - | - |
| 12 | ND | ND | - | - | - | - | - | - |
| 13 | 32,3 | 31,5 | 31,9 | 0,573 | 1455,80 | 2477,82 | 1966,81 | 722,67 |
| 14 | ND | ND | - | - | - | - | - | - |
| 15 | ND | ND | - | - | - | - | - | - |
| 16 | ND | ND | - | - | - | - | - | - |
| 17 | 28,0 | 27,2 | 27,6 | 0,622 | 23525,70 | 41924,64 | 32725,17 | 13010,01 |
| 18 | ND | ND | - | - | - | - | - | - |
| 19 | 31,4 | 30,8 | 31,1 | 0,467 | 2540,41 | 3918,32 | 3229,37 | 974,32 |
| 20 | ND | ND | - | - | - | - | - | - |
| 21 | ND | ND | - | - | - | - | - | - |
| 22 | ND | ND | - | - | - | - | - | - |
| 23 | ND | ND | - | - | - | - | - | - |
| 24 | 30,7 | 30,0 | 30,3 | 0,495 | 4129,63 | 6539,01 | 5334,32 | 1703,69 |
| 25 | 30,5 | 29,8 | 30,2 | 0,552 | 4527,22 | 7555,16 | 6041,19 | 2141,08 |
| 26 | ND | ND | - | - | - | - | - | - |
| 27 | 31,6 | 31,1 | 31,4 | 0,410 | 2198,73 | 3217,78 | 2708,26 | 720,57 |
| 28 | 33,0 | 32,3 | 32,6 | 0,495 | 912,18 | 1444,38 | 1178,28 | 376,32 |
| 29 | 35,1 | 35,6 | 35,3 | 0,339 | 226,76 | 165,46 | 196,11 | 43,35 |
| 30 | 31,2 | 32,2 | 31,7 | 0,707 | 2935,19 | 1522,27 | 2228,73 | 999,08 |
| 31 | 33,9 | 34,3 | 34,1 | 0,332 | 508,51 | 373,49 | 441,00 | 95,47 |
| 32 | ND | ND | - | - | - | - | - | - |
| 33 | 38,1 | 38,4 | 38,2 | 0,177 | 31,22 | 26,49 | 28,86 | 3,34 |
| 34 | 42,2 | 40,2 | 41,2 | 1,386 | 2,21 | 8,02 | 5,12 | 4,11 |
| 35 | 40,5 | 41,0 | 40,8 | 0,368 | 6,54 | 4,65 | 5,60 | 1,34 |
| 36 | ND | ND | - | - | - | - | - | - |
| 37 | ND | ND | - | - | - | - | - | - |
| 38 | ND | ND | - | - | - | - | - | - |
| 39 | 41,0 | 42,6 | 41,8 | 1,089 | 4,68 | 1,70 | 3,19 | 2,11 |
| 40 | 39,1 | 40,1 | 39,6 | 0,686 | 16,41 | 8,68 | 12,54 | 5,46 |
| 41 | 29,1 | 28,9 | 29,0 | 0,120 | 11576,76 | 12943,79 | 12260,28 | 966,64 |
| 42 | 36,8 | 37,4 | 37,1 | 0,424 | 76,75 | 51,76 | 64,26 | 17,67 |
| 43 | 40,1 | 39,5 | 39,8 | 0,410 | 8,51 | 12,45 | 10,48 | 2,79 |
| 44 | 31,1 | 31,2 | 31,1 | 0,099 | 3238,98 | 2954,52 | 3096,75 | 201,14 |
| 45 | ND | ND | - | - | - | - | - | - |
| 46 | ND | ND | - | - | - | - | - | - |
| 47 | ND | ND | - | - | - | - | - | - |
| 48 | 42,8 | 43,8 | 43,3 | 0,707 | 1,49 | 0,77 | 1,13 | 0,51 |
| 49 | 42,2 | 42,9 | 42,5 | 0,4949747 | 2,19 | 1,38 | 1,78 | 0,57 |
| 50 | 42,5 | 44,2 | 43,3 | 1,2091526 | 1,77 | 0,58 | 1,17 | 0,85 |
| 51 | 38,1 | 38,4 | 38,3 | 0,1979899 | 31,43 | 26,15 | 28,79 | 3,73 |
| 52 | ND | ND | - | - | - | - | - | - |
| 53 | ND | ND | - | - | - | - | - | - |
| 54 | 40,1 | 42,9 | 41,5 | 1,9940411 | 8,62 | 1,35 | 4,99 | 5,14 |
| 55 | ND | ND | - | - | - | - | - | - |
| 56 | 42,6 | 41,5 | 42,1 | 0,7212489 | 1,69 | 3,31 | 2,50 | 1,14 |
| 57 | ND | ND | - | - | - | - | - | - |
| 58 | ND | ND | - | - | - | - | - | - |
| 59 | ND | ND | - | - | - | - | - | - |
| 60 | ND | ND | - | - | - | - | - | - |
| 61 | ND | ND | - | - | - | - | - | - |
| 62 | ND | ND | - | - | - | - | - | - |
| 63 | ND | ND | - | - | - | - | - | - |
| 64 | ND | ND | - | - | - | - | - | - |
| 65 | ND | ND | - | - | - | - | - | - |
| 66 | ND | ND | - | - | - | - | - | - |
| 67 | ND | ND | - | - | - | - | - | - |
| 68 | ND | ND | - | - | - | - | - | - |
| 69 | ND | ND | - | - | - | - | - | - |
| 70 | ND | ND | - | - | - | - | - | - |
| 71 | ND | ND | - | - | - | - | - | - |
| 72 | ND | ND | - | - | - | - | - | - |

Cq= Ct= threshold cycle

VL= Viral load, calculated using the estándar curve equation.

SD= Standar deviation

**Supplemtary 1. Cq values of each sample and viral load (Copies / mL)**

| **Sample number** | | | | | |
| --- | --- | --- | --- | --- | --- |
| **ID** | **April 2021** | **June 2021** | **September 2021** | **December 2021** | **March 2021** |
| SP1 | 1 | 21 | 37 | 45 | 61 |
| SP2 | 2 | 22 | 38 | 46 | 62 |
| SP3 | 3 | 23 | 39 | 47 | 63 |
| SP4 | 4 | 19 | 35 | 43 | 59 |
| SP5 | 5 | 20 | 36 | 44 | 60 |
| SP6 | 6 | 18 | 34 | 42 | 58 |
| SP7 | 7 | 24 | 40 | 48 | 64 |
| SP8 | 8 | 17 | 33 | 41 | 65 |
| SP9 | 9 | 25 |  | 49 | 57 |
| SP10 | 10 | 27 |  | 51 | 67 |
| SP11 | 11 | 32 |  | 56 | 72 |
| SP12 | 12 | 31 |  | 55 | 71 |
| SP13 | 13 | 30 |  | 54 | 70 |
| SP14 | 14 | 29 |  | 53 | 69 |
| SP15 | 15 | 28 |  | 52 | 68 |
| SP16 | 16 | 26 |  | 50 | 66 |

Supplementary 2.
